# Pain Trajectories and Mechanisms in Early Inflammatory Arthritis: Results from a 2-Year Prospective Cohort

**DOI:** 10.64898/2026.01.08.26343656

**Authors:** Zoe Rutter-Locher, Sam Norton, Bina Menon, Tom Esterine, Ruth Williams, Leonie S. Taams, Kirsty Bannister, Bruce W. Kirkham

## Abstract

**Introduction:** Pain is often persistent in patients with inflammatory arthritis (IA). There is limited research on pain mechanisms in early IA. We used detailed assessments to characterise pain types longitudinally.

**Methods:** This prospective observational study investigated the relative contribution of peripheral versus centrally mediated pain in newly diagnosed IA patients with pain scores ≥3, followed over two years. Assessments included: disease activity (Disease Activity Score-28, musculoskeletal ultrasound); quality of life (Musculoskeletal Health questionnaire); mental health status (Patient Health Questionnaire Anxiety, Depression Scale) and pain characteristics (Fibromyalgia criteria, painDETECT, Static and Dynamic Quantitative Sensory Testing). Mixed-effects regression models examined longitudinal associations.

**Results:** Among 66 participants (all baseline pain NRS ≥3), pain decreased significantly in the first six months, with smaller, non-significant reductions thereafter. At 24 months, 49% still reported pain (NRS ≥3) and 25% showed no pain improvement. Those with pain had higher baseline and longitudinal scores for centrally mediated pain (fibromyalgia severity, painDETECT) and psychological distress, despite similar baseline inflammation and similar reductions over time.

Both baseline and longitudinal inflammation was not associated with subsequent pain (*b* –0.05, 95% CI –0.67 to 0.56), whereas higher centrally mediated pain markers predicted less pain reduction (*b* 1.41, 95% CI 0.83–1.99, p<0.001).

**Conclusion:** In our early IA cohort, despite reductions in inflammation half of patients reported persistent pain, primarily driven by centrally mediated mechanisms. Early identification and treatment of this group may improve long-term outcomes.

**Key messages:**

- This is the first longitudinal study assessing relative contributions of inflammation and centrally mediated pain features in early IA.
- Despite reductions in inflammation half of patients reported persistent pain, primarily driven by centrally mediated mechanisms.
- Early assessment and treatment of non-inflammatory pain mechanisms may improve long-term pain outcomes.

## Introduction

Advances in anti-inflammatory therapies have led to major improvements in outcomes for many patients with inflammatory arthritis (IA). However, it is increasingly recognised that up to 30% of patients have persistent, unacceptable pain [1–4]. Pain is consistently prioritised by patients as a major concern, often above other symptoms such as fatigue, swelling or stiffness [5–8]. It is strongly associated with reduced quality of life, physical disability, and reduced ability to participate in social activities[9] and the workforce[10]. Persistent pain is also closely linked to psychological distress, including depression and anxiety, which may be a risk factor for or a consequence of persistent pain, and is associated with poorer treatment adherence and outcomes[11,12]. Despite the increased recognition of pain as a major problem in IA, very little research has been done in the early stages of IA. Our recent work highlighted that altered pain processing, including increased pain sensitivity and features of centrally mediated pain, may already be present at diagnosis, as identified through clinical evaluation, patient-reported outcomes, musculoskeletal ultrasound, and quantitative sensory testing[13].

In established IA, persistent pain is increasingly recognised as arising from a complex interplay between ongoing inflammatory and non-inflammatory mechanisms, including peripherally and centrally mediated pain due to maladaptive nociceptive processing[14,15]. These different drivers of pain are often acting simultaneously requiring detailed individual assessment to accurately identify causes of pain.

Failure to understand the mechanistic underpinning of a patients’ pain may lead to ineffective approaches to analgesia, including increased opioid prescribing and inappropriate escalation of immunosuppressive therapy, adding burden to healthcare systems[16].

Here, we present a comprehensive, longitudinal analysis of pain processing mechanisms in a real-world, early inflammatory arthritis cohort. Building on our previously reported baseline findings at diagnosis[17], we follow the same patients over a two-year period to prospectively investigate the evolving contributions of inflammatory versus non-inflammatory, and likely peripherally versus centrally-mediated mechanisms of pain and identify clinical, psychological, and sensory predictors of persistent pain.

## Methods

### Study Design and Ethical Approval

The **“**Pain phenotypes and their Underlying Mechanisms in Inflammatory Arthritis” (PUMIA) study is a single-site, prospective observational study designed to investigate pain mechanisms in patients with IA. Patients with new onset IA were recruited soon after diagnosis and followed up at 4 timepoints over 2 years. All participants provided written informed consent, and the study received ethical approval from the Bromley Research Ethics Committee and the Health Research Authority (REC reference: 21/LO/0712).

### Participant Recruitment and Assessments

Individuals who reported a total numerical rating scale (NRS) pain score of 3 or higher on a scale of 0 to 10, received a clinician diagnosis of peripheral inflammatory joint disease, and experienced symptom onset within the past 12 months were recruited. Patients were enrolled in the study as soon as feasible following their diagnosis. Full recruitment criteria are available in supplementary material (Supplementary Data S1) and assessment protocols, and quantitative sensory testing (QST) procedures, have been published previously[17]. At the baseline study visit (at diagnosis) and 1 year, participants underwent full in-person assessments, which included (i) Pain and quality of life assessment: pain ratings on a 0–10 numerical rating scale (NRS), of both overall pain and pain in their most affected joint, in the previous 24 hours and the Musculoskeletal Health Questionnaire (MSK-HQ) for quality of life (ii) Clinical inflammatory disease activity assessment: High resolution musculoskeletal ultrasonography (HRUS-14 joints), CRP, 28-TJC, 28-SJC, DAS28-CRP, (iii) Assessment to identify mechanisms of pain that are more likely to be centrally mediated: patient reported outcomes measures (PROMS) including the Patient Health Questionnaire-9 (PHQ-9) for depression, the Generalised Anxiety Disorder-7 (GAD-7) for anxiety, the Patient Health Questionnaire-15 (PHQ-15) for somatic symptom burden, the 2016 revision of the American College of Rheumatology (ACR) 2010 modified diagnostic criteria for fibromyalgia ( comprising the Widespread Pain Index (WPI) and the Symptom Severity Scale (SSS), fibromyalgia severity was calculated as the sum of the WPI and SSS scores), and the painDETECT questionnaire (a validated screening tool for neuropathic-like pain, commonly used as a proxy for identifying centrally mediated pain due to the similarity in symptom profiles) alongside static and dynamic QST according to previously reported protocols[13]. At 6 months and 2 years, participants completed remote follow-up consisting of pain ratings and PROMs (MSK-HQ, PHQ-9, GAD-7, PHQ-15, fibromyalgia criteria and severity, and painDETECT).

### Sample size

Our sample size was based on our original hypothesis that peripheral inflammatory pain would predominate at diagnosis, but that, over disease course, centrally mediated pain mechanisms would develop and contribute to the pain state in some people. A target sample size of 60 was calculated based on providing acceptable balance between feasibility of recruitment and precision in the estimate of the proportion of people transitioning from a peripheral pain phenotype to a centrally mediated pain phenotype (expected to be 30% based on literature), where the maximum width of the 95% CI was ±12.8%. Although relatively wide, we considered this a feasible number to recruit and acceptable when considered alongside the use of continuous scores to provide additional contextual information in addition to the dichotomised variable.

### Statistical analysis

All data are presented as mean ± standard deviation (SD). To normalise the data for MDT and MPT, logarithmic transformations were applied. Absolute reference data (matched for age, gender and site, being the dorsum of the hand) was used to normalise test results of the individual participant by calculating the z transformation= (value participant-mean controls))/SD controls (29). A z-score of beyond ±1.96 is considered to reflect values in the extreme 5% of the healthy reference distribution (i.e. 2.5% in each tail) corresponded to statistical significance at the 0.05% level .

#### Pain trajectories

Mixed-effects linear regression models were used to assess changes in continuous outcomes (e.g. pain, inflammatory markers, mental health, centrally mediated pain markers, and quality of life) over time, with a random intercept to account for repeated measures within participants over time. Time was included as a dummy coded categorical fixed effect allowing for the estimation of timepoint specific marginal means with 95% confidence intervals based on a missing at random assumption. Pairwise comparisons between time points were performed with Bonferroni correction. For binary outcomes (e.g. proportion reporting pain ≥3), mixed-effects logistic regression was used, with predicted probabilities over time obtained from the model. To enable direct comparison of change over time across variables measured on different scales, all continuous variables were standardised to baseline values. For each variable, the mean and SD at baseline were calculated, and z-scores were computed at each subsequent timepoint as (x-baseline mean) /baseline SD.

Associations between inflammation-related and centrally mediated pain–related factor scores and residualised change in pain were examined using separate linear mixed-effects models with random intercepts for participants, with pain as the outcome variable. These models controlled for inflammation or centrally mediated pain factor scores at the previous time point, prior pain, age, gender, and time. Factor scores were derived via exploratory factor analysis (principal factoring) : One factor was extracted from inflammation-related variables (DAS28-Swollen Joint Count, C-reactive protein, Evaluator Global Assessment, HRUS-PD score) and another from centrally mediated pain-related variables ( painDETECT, fibromyalgia severity, PHQ-9 and pressure pain threshold at right trapezius).

#### Predictors of persistent pain

Two-sided independent t-tests were used to compare baseline differences between participants with and without clinically relevant pain (NRS ≥3) at 6, 12, and 24 months. For each variable, we report the mean difference (95% CI) and effect sizes using Cohen’s *d* (95% CI). Effect sizes were interpreted using standard thresholds: 0.2 (small), 0.5 (medium), and 0.8 (large).

Exploratory mixed-effects linear regression models were then used to examine whether baseline predictors (factor-derived central and inflammatory scores, as well as individual baseline variables) were associated with subsequent pain, accounting for repeated measures and including time-by-predictor interaction terms.

For all analyses a p-value <0.05 was considered statistically significant. Analyses were conducted in Stata V17.0 (StataCorp, College Station, Texas, USA) and R Studio using R version 4.5.0 (2025-04-11)

## Results

### Study Cohort and Follow-Up Completion

66 participants were recruited, with a mean age of 50 years, 56% female and diverse ethnicity (Table 1). The majority were diagnosed with rheumatoid arthritis (62%). Mean time since diagnosis was 1.2 months. At baseline, most patients had active disease (mean DAS28-CRP 3.9) and active synovitis on examination +/- MSK US (65 with ≥1 active joint). Mean overall pain score was 5.4/10, and 67% and 62% screened positive for depression and anxiety, respectively. 20-30% of patients fulfilled criteria for widespread pain, fibromyalgia and likely neuropathic like pain on painDETECT questionnaire. On QST, as a group there were no abnormalities in skin sensitivity (all z scores < 1.96) and PPT were lower at joint sites than at trapezius. QST markers supported the evidence of centrally mediated pain, with 16% fulfilling criteria for TSP and 61% non-responders to CPM PDT paradigm (Table 1).

**Table 1.**
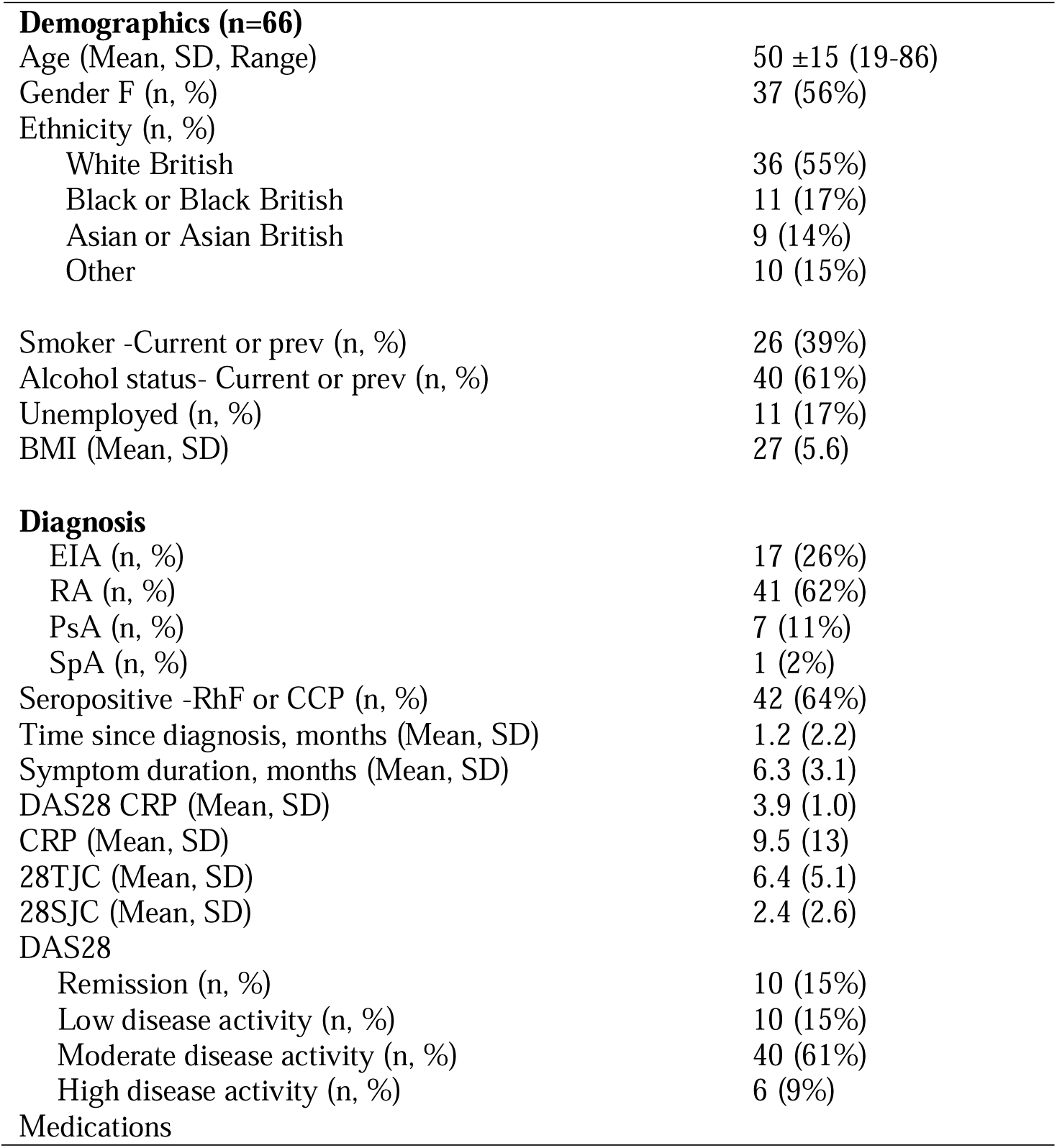
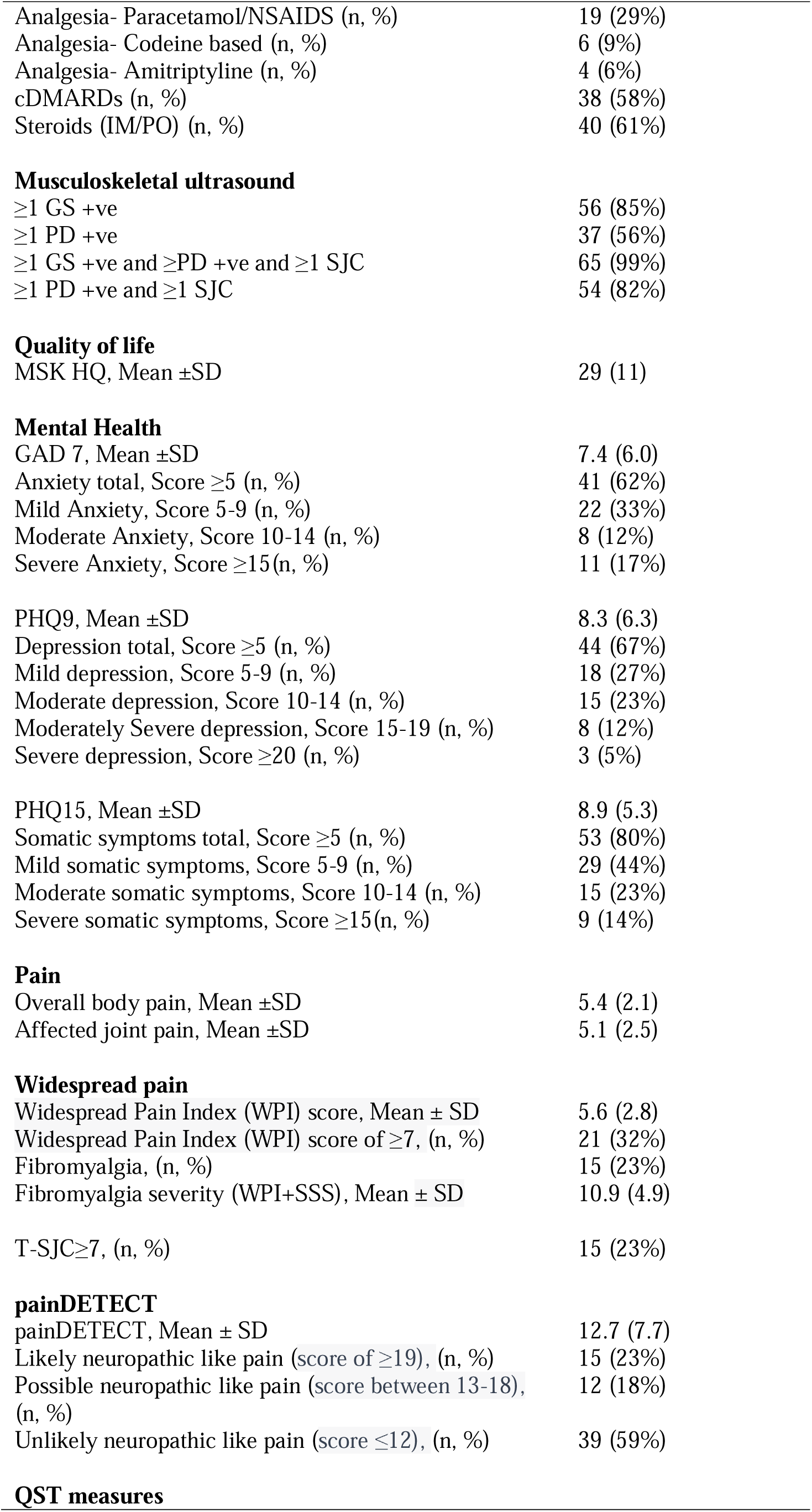
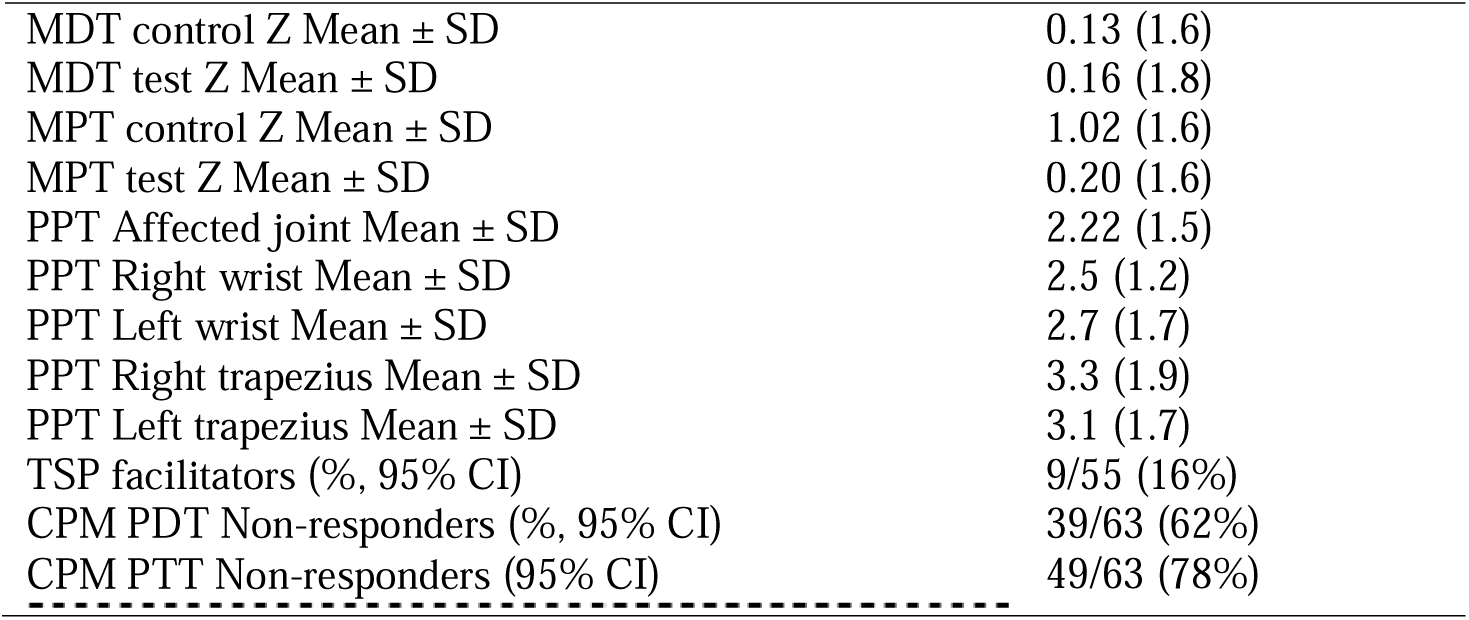
Patient demographics, diagnosis and medication alongside results of questionnaire data collected for quality of life, mental health, fibromyalgia criteria and painDETECT. (n=66). Abbreviations: BMI (Body mass index), EIA (Early inflammatory arthritis), RA (Rheumatoid arthritis), PsA (Psoriatic arthritis), SpA (Spondylarthritis), RhF (Rheumatoid factor), CCP (cyclic citrullinated peptide), IM (Intra-muscular), PO (Oral), IV (Intra-venous), DMARDS (Disease modifying anti-rheumatic drugs), MSK HQ (Musculoskeletal health questionnaire), GAD 7 (Generalised anxiety disorder 7 questionnaire), PHQ9 (Patient health questionnaire 9), PHQ15 (Patient health questionnaire 15).

**Demographics (n=66)**
|  |  |
| --- | --- |
| Age (Mean, SD, Range) | 50 ±15 (19-86) |
| Gender F (n, %) | 37 (56%) |
| Ethnicity (n, %) |  |
| White British | 36 (55%) |
| Black or Black British | 11 (17%) |
| Asian or Asian British | 9 (14%) |
| Other | 10 (15%) |
| Smoker -Current or prev (n, %) | 26 (39%) |
| Alcohol status- Current or prev (n, %) | 40 (61%) |
| Unemployed (n, %) | 11 (17%) |
| BMI (Mean, SD) | 27 (5.6) |
| **Diagnosis** |  |
| EIA (n, %) | 17 (26%) |
| RA (n, %) | 41 (62%) |
| PsA (n, %) | 7 (11%) |
| SpA (n, %) | 1 (2%) |
| Seropositive -RhF or CCP (n, %) | 42 (64%) |
| Time since diagnosis, months (Mean, SD) | 1.2 (2.2) |
| Symptom duration, months (Mean, SD) | 6.3 (3.1) |
| DAS28 CRP (Mean, SD) | 3.9 (1.0) |
| CRP (Mean, SD) | 9.5 (13) |
| 28TJC (Mean, SD) | 6.4 (5.1) |
| 28SJC (Mean, SD) | 2.4 (2.6) |
| DAS28 |  |
| Remission (n, %) | 10 (15%) |
| Low disease activity (n, %) | 10 (15%) |
| Moderate disease activity (n, %) | 40 (61%) |
| High disease activity (n, %) | 6 (9%) |

**Table 1** Patient demographics, diagnosis and medication alongside results of questionnaire data collected for quality of life, mental health, fibromyalgia criteria and painDETECT.
|  |  |
| --- | --- |
| Analgesia- Paracetamol/NSAIDS (n, %) | 19 (29%) |
| Analgesia- Codeine based (n, %) | 6 (9%) |
| Analgesia- Amitriptyline (n, %) | 4 (6%) |
| cDMARDs (n, %) | 38 (58%) |
| Steroids (IM/PO) (n, %) | 40 (61%) |
| **Musculoskeletal ultrasound** |  |
| ≥1 GS +ve | 56 (85%) |
| ≥1 PD +ve | 37 (56%) |
| ≥1 GS +ve and ≥PD +ve and ≥1 SJC | 65 (99%) |
| ≥1 PD +ve and ≥1 SJC | 54 (82%) |
| **Quality of life** |  |
| MSK HQ, Mean ±SD | 29 (11) |
| **Mental Health** |  |
| GAD 7, Mean ±SD | 7.4 (6.0) |
| Anxiety total, Score ≥5 (n, %) | 41 (62%) |
| Mild Anxiety, Score 5-9 (n, %) | 22 (33%) |
| Moderate Anxiety, Score 10-14 (n, %) | 8 (12%) |
| Severe Anxiety, Score ≥15(n, %) | 11 (17%) |
| PHQ9, Mean ±SD | 8.3 (6.3) |
| Depression total, Score ≥5 (n, %) | 44 (67%) |
| Mild depression, Score 5-9 (n, %) | 18 (27%) |
| Moderate depression, Score 10-14 (n, %) | 15 (23%) |
| Moderately Severe depression, Score 15-19 (n, %) | 8 (12%) |
| Severe depression, Score ≥20 (n, %) | 3 (5%) |
| PHQ15, Mean ±SD | 8.9 (5.3) |
| Somatic symptoms total, Score ≥5 (n, %) | 53 (80%) |
| Mild somatic symptoms, Score 5-9 (n, %) | 29 (44%) |
| Moderate somatic symptoms, Score 10-14 (n, %) | 15 (23%) |
| Severe somatic symptoms, Score ≥15(n, %) | 9 (14%) |
| **Pain** |  |
| Overall body pain, Mean ±SD | 5.4 (2.1) |
| Affected joint pain, Mean ±SD | 5.1 (2.5) |
| **Widespread pain** |  |
| Widespread Pain Index (WPI) score, Mean ± SD | 5.6 (2.8) |
| Widespread Pain Index (WPI) score of ≥7, (n, %) | 21 (32%) |
| Fibromyalgia, (n, %) | 15 (23%) |
| Fibromyalgia severity (WPI+SSS), Mean ± SD | 10.9 (4.9) |
| T-SJC≥7, (n, %) | 15 (23%) |
| **painDETECT** |  |
| painDETECT, Mean ± SD | 12.7 (7.7) |
| Likely neuropathic like pain (score of ≥19), (n, %) | 15 (23%) |
| Possible neuropathic like pain (score between 13-18), (n, %) | 12 (18%) |
| Unlikely neuropathic like pain (score ≤12), (n, %) | 39 (59%) |
| **QST measures** |  |
| MDT control Z Mean ± SD | 0.13 (1.6) |
| MDT test Z Mean ± SD | 0.16 (1.8) |
| MPT control Z Mean ± SD | 1.02 (1.6) |
| MPT test Z Mean ± SD | 0.20 (1.6) |
| PPT Affected joint Mean ± SD | 2.22 (1.5) |
| PPT Right wrist Mean ± SD | 2.5 (1.2) |
| PPT Left wrist Mean ± SD | 2.7 (1.7) |
| PPT Right trapezius Mean ± SD | 3.3 (1.9) |
| PPT Left trapezius Mean ± SD | 3.1 (1.7) |
| TSP facilitators (% , 95% CI) | 9/55 (16%) |
| CPM PDT Non-responders (% , 95% CI) | 39/63 (62%) |
| CPM PTT Non-responders (95% CI) | 49/63 (78%) |

Follow-up rates were acceptable across the study period, with 61 (92%) of participants completing at least one follow-up assessment. Data was available for 53 participants (80%) at 6 months, 58 (88%) at 12 months, and 57 (86%) at 24 months.

### Pain Trajectories Over 2 years

Over the study period, mean overall pain score reduced from 5.4 (SD 2.1) at baseline, to 4.1 (2.8) at 6 months, 3.7 (2.8) at 12 months and 3.1 (3.1) at 24 months. Using a mixed effect linear regression model, there was a significant reduction in pain between baseline and 6 months (p<0.01), with further gradual decreases between subsequent consecutive time points that did not reach statistical significance.

At 24 months 49% (95% CI: 36–61%) of participants reported persistent overall pain ≥3 (Figure 1). Of note, 25% of the participants had pain that did not change or worsened over the 2-year period. Pain score changes showed considerable individual variation (mean -2.2, SD 3.1; range -9 to +7)(Supplementary Figure S1).

**Figure 1.**
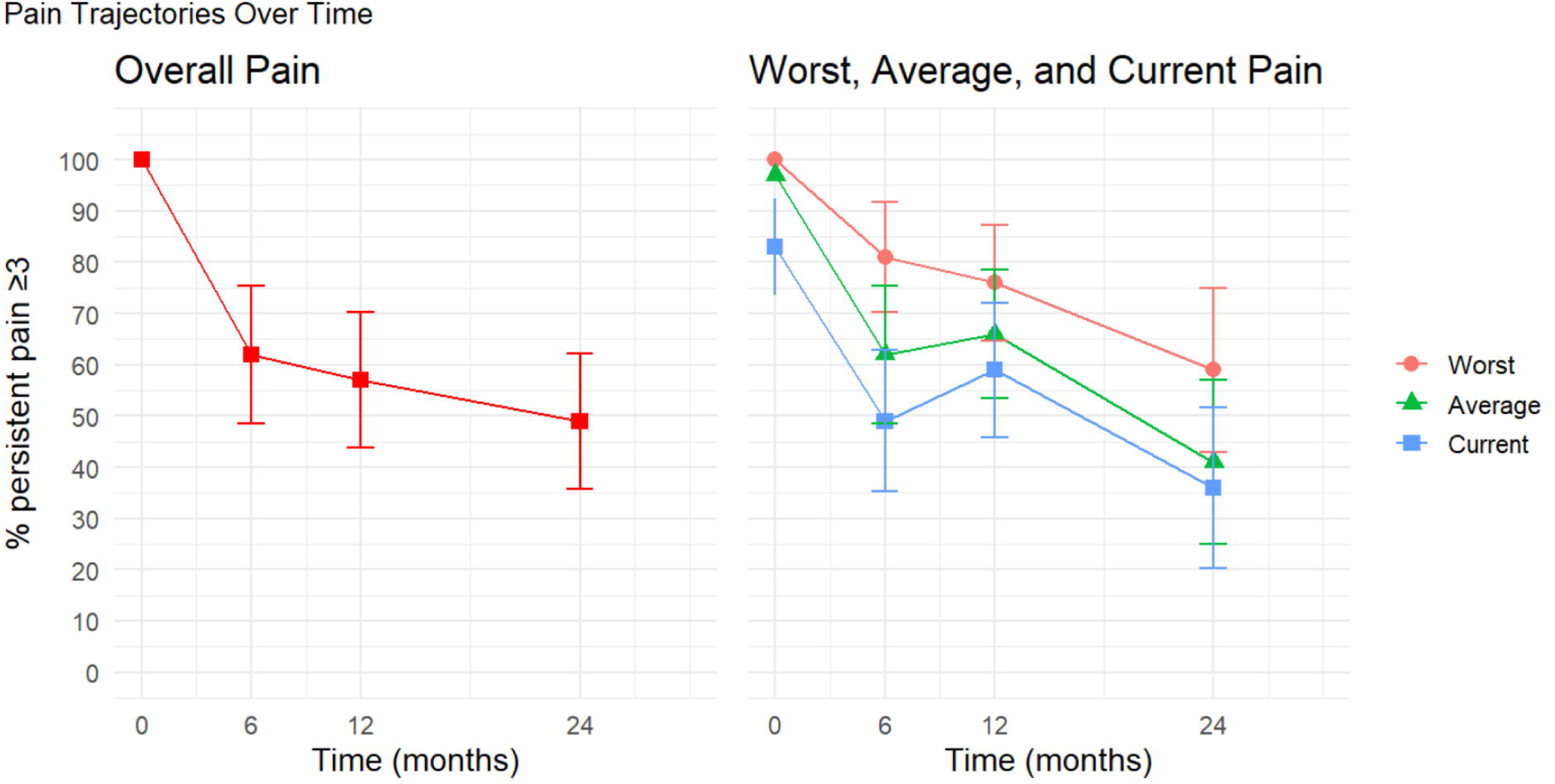
Pain Trajectories Over Time in Early Inflammatory Arthritis. Figure shows the percentage of participants reporting persistent pain (score ≥3 on 0–10 numerical rating scale) over 24 months. Left panel shows overall pain; right panel shows worst, average, and current pain scores at each time point, from painDETECT questionnaire. Although overall pain improved, 49% of patients continued to report clinically relevant pain. Total N at each time point: baseline (n=66), 6 months (n=53), 12 months (n=58), 24 months (n=57). Error bars represent 95% confidence intervals.

On painDETECT, 36% (95% CI: 25–49%) reported current pain ≥3, 41% (95% CI: 29–54%) average pain ≥3, and 59% (95% CI: 46–71%) worst pain was ≥3 over the previous 4 weeks (Figure 1). Using a mixed-effects logistic regression model, the likelihood of overall pain ≥3 declined over time: 64% at 6 months (95% CI: 52–77%), 57% at 12 months (95% CI: 44–70%), and 50% at 24 months (95% CI: 37–63%), though this trend did not reach statistical significance (p=0.17).

### Trajectories of Inflammatory and Centrally mediated Pain Markers

Over 24 months, observed raw means (Supplementary Table S1) and modelled estimates (Supplementary Table S2, Figure 2) showed significant improvement in PROM markers of centrally mediated pain (Modelled estimates: fibromyalgia severity –2.71; painDETECT –2.71), mental health (GAD-7 –2.31; PHQ-9 –1.66; PHQ-15 –1.93) and inflammation (CRP –4.90; DAS28-SJC –1.70; EGA –12.60). MSK-HQ scores improved significantly (+10.29), more than the minimal clinically important difference[18]. Fatigue showed a non-significant reduction (–4.37).

**Figure 2:**
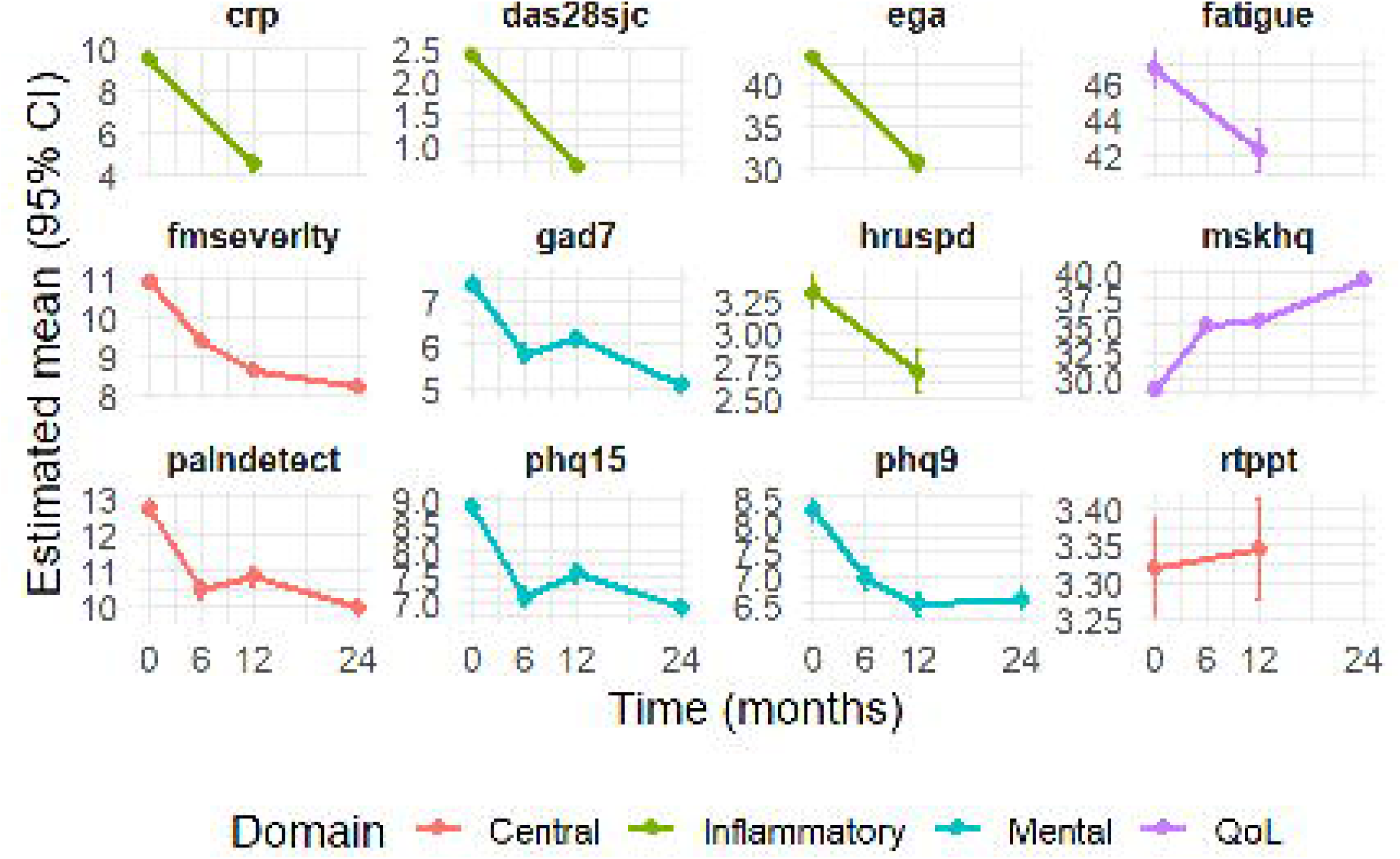
Modelled trajectories of centrally mediated pain, mental health and inflammatory measures over 24 months. Graphs show **estimated marginal means (±95% confidence intervals) for 12 measures across four domains: Centrally mediated pain (orange), Mental health (blue), Inflammation (green), and Quality of life (purple). Results are derived from mixed-effects linear regression models with time as a fixed effect and participant ID as a random intercept. Each panel shows predicted outcome values at 0, 6, 12, and 24 months. Markers of centrally mediated pain reduced over time: PainDETECT** scores significantly decreased from 12.7 at baseline to 10.0 at 24 months, with modelled reductions at all follow-up points compared to baseline (*p* = 0.014 at 24 months). **Fibromyalgia Severity Scores** fell from 10.9 at baseline to 8.2 at 24 months, with significant reductions at all time points (*p* = 0.031 to *p* < 0.001).**Trapezius PPT** z-scores remained stable over time, with no significant change between baseline and 12 months (*p* = 0.904).**Mental health scores reduced over time: PHQ-9** reduced from 8.3 to 6.6, with significant reductions at 12 and 24 months (p = 0.016 and p = 0.034, respectively). **GAD-7** decreased from 7.4 at baseline to 5.1 at 24 months, with significant reductions at 6 months (*p* = 0.021) and 24 months (*p* < 0.001). **PHQ-15** reduced from 8.9 to 6.9, with all timepoints significantly lower than baseline (all *p* < 0.05). Inflammatory markers reduced: **CRP** levels reduced from 9.5Dmg/L at baseline to 4.6Dmg/L at 12 months (p = 0.001). **DAS28-SJC** decreased from 2.4 to 0.7 (p < 0.001), and **EGA** from 43.2 to 30.6 (p < 0.001). **MSK ultrasound (HRUS PD)** scores reduced from 9.1 to 7.3, though this was not statistically significant (p = 0.096). **Quality of life (QoL)** measures improved with MSK-HQ scores improved significantly from 29.0 at baseline to 39.3 at 24 months (p < 0.001). However, **fatigue** scores showed a small, non-significant reduction from 46.7 to 42.3 (p = 0.30). CRP: C-reactive protein DAS28sjc: Disease Activity Score Swollen Joint Count (28 joints); EGA: Evaluator Global Assessment; FM: Fibromyalgia; GAD-7: Generalised Anxiety Disorder Scale; HRUS: High-Resolution Ultrasound; MSK-HQ: Musculoskeletal Health Questionnaire; PHQ-9: Patient Health Questionnaire–Depression; PHQ-15: Patient Health Questionnaire–Somatic Symptoms; rttppt: Pressure Pain Threshold at Right trapezius

Modelled estimates showed significant increase in joint PPT, indicating a reduction in peripheral joint sensitivity (Target joint PPT 0.72, right wrist 0.87, left wrist 0.64) but no significant difference in trapezius PPT (right trapezius 0.02, left trapezius -0.002). There were no significant differences in TSP or CPM effects (TSP 0.95, CPM PDT effect 2.13, CPM PTT effect -0.86) (Supplementary Table S2)

Using z scores, which allows direct comparison of trends of different measures, fibromyalgia severity, SJC and evaluator global assessment (EGA) showed the greatest reduction over time on a standardised scale (Supplementary Figure S2).

### Association Between Pain Reduction and Peripheral Inflammatory vs Centrally mediated Pain Mechanisms

In a mixed effects regression model, using factor-derived scores and modelling residualised change in pain (i.e. controlling for prior pain) over 2 years, inflammation at the previous time point was not associated with subsequent pain (*b*= –0.05, 95% CI –0.67 to 0.56, p = 0.87) but centrally mediated pain at the previous time point was strongly and consistently associated with higher subsequent pain relative to expected pain given the prior level (*b* = 1.41, 95% CI 0.83 to 1.99, *p* < 0.001), meaning centrally mediated pain measures were associated with reduced improvement in pain .

These relationships were observed across all time points (Inflammation: 6 months: *b* = 0.20, 95% CI – 0.59 to 0.99, p = 0.62; 24 months: *b* = –0.34, 95% CI –1.11 to 0.44, p = 0.39, Central: 6 months: *b* = 1.59, 95% CI 0.83 to 2.36, *p* < 0.001; 12 months: *b* = 1.23, 95% CI 0.44 to 2.02, *p* = 0.002; 24 months: *b* = 1.39, 95% CI 0.59 to 2.20, *p* = 0.001).

### Baseline Predictors of Persistent Pain

Individuals with pain ≥3 at 6, 12, and 24 months consistently had higher baseline centrally mediated pain markers (fatigue, patient global assessment (PGA), painDETECT scores, fibromyalgia severity (at 6 and 24 months), and worse mental health and quality of life (MSKHQ))(Supplementary Table S3). The largest effect sizes were for fatigue, GAD7, PHQ15 and painDETECT. There were no consistent significant differences between those with or without pain ≥3 in demographics, symptom duration, pain or TJC, inflammatory markers, or QST measures at baseline (Supplementary Table S3). Those individuals with persistent pain also had consistently higher levels of centrally mediated pain markers across all timepoints, despite similar baseline inflammation and comparable reductions in inflammation over time (Figure 3). Within the group of individuals with persistent pain at 24 months (28, 49%) , there was a trend for two pain trajectories; pain that reduced from baseline but was still ≥3 (14, 25%) and pain that did not change or worsened (14, 25%)(Supplementary Figure S3).

**Figure 3.**
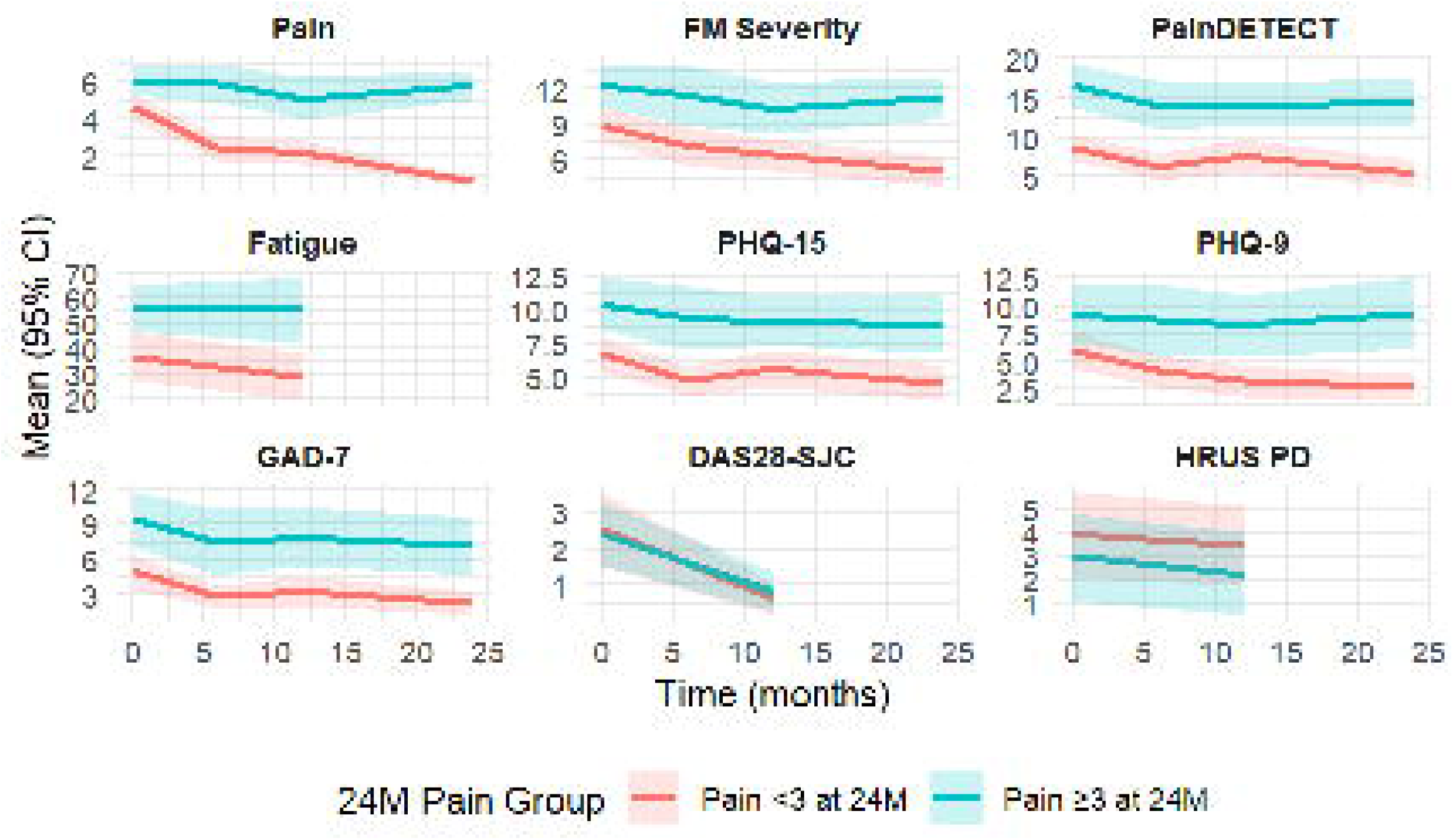
Trajectories of clinical measures, stratified by 24-month pain outcome. Graphs show mean scores (±95% CI) over time for participants with low pain (NRS <3) vs. persistent pain (NRS ≥3) at 24 months. Those in pain at 24months had consistently reported pain over disease course, but those without pain had reduced their pain from baseline. The persistent pain group had consistently higher levels of FM Severity, PainDETECT scores, psychological distress and fatigue across all timepoints. Measures of inflammation (DAS28-SJC, HRUS PD score) reduced comparably in both groups.

#### Factor scores

In a mixed effects regression model, using factor-derived scores, baseline inflammation was not associated with subsequent pain (*b* = 0.05, 95% CI –0.50 to 0.59, p = 0.87) whereas baseline centrally mediated pain was strongly and consistently associated with higher subsequent pain (*b* = 1.32, 95% CI 0.83 to 1.82, p < 0.001).

These relationships were observed across all time points (Inflammation: 6 months: *b* = 0.24, 95% CI – 0.56 to 1.04, p = 0.56; 12 months: *b* = –0.19, 95% CI –0.94 to 0.56, p = 0.62; 24 months: *b* = –0.37, 95% CI –1.12 to 0.38, p = 0.33. Central: baseline: *b* = 0.92, 95% CI 0.25 to 1.59, p = 0.007; 6 months: *b* = 1.81, 95% CI 1.04 to 2.58, p < 0.001; 12 months: *b* = 1.19, 95% CI 0.47 to 1.92, p = 0.001; 24 months: *b* = 1.46, 95% CI 0.73 to 2.19, p < 0.001).

These findings suggest that baseline centrally mediated pain variables, rather than inflammation, are strongly associated with persistent pain.

#### Individual variables

To explore which individual centrally mediated pain and psychological distress variables may be more strongly associated with persistent pain, the mixed-effect regression model was repeated using standardised individual centrally mediated pain variables. Psychological and PROMs of centrally mediated pain (PHQ-15, fatigue, GAD-7, PHQ-9, painDetect, and fibromyalgia severity) were the strongest predictors of subsequent pain, each showing large and consistent associations. Tender-swollen joint count difference showed a smaller but significant effect. Quantitative sensory testing measures, including trapezius PPT, TSP and CPM, were not significantly associated with later pain (Table 2).

**Table 2.** Standardised baseline predictors of subsequent pain. Results from mixed-effects linear regression models showing average marginal effects (AME) of each predictor on pain, expressed per 1 SD increase. Models adjusted for age and gender, with repeated measures accounted for by random intercepts. Positive AMEs indicate higher subsequent pain, whereas negative AMEs indicate lower subsequent pain. PHQ15 (Patient health questionnaire 15), GAD 7 (Generalised anxiety disorder 7 questionnaire), PHQ9 (Patient health questionnaire 9), FM (Fibromyalgia) , TJC-SJC diff (Tender minus swollen joint count 28 joints), rTPPT (Right trapezius pressure pain threshold), CPM (Conditioned pain modulation), TSP (Temporal summation of pain).

| Predictor | AME (per 1 SD) | 95% CI | p-value |
| --- | --- | --- | --- |
| **PHQ-15** | 1.23 | 0.78 – 1.69 | <0.001 |
| **Fatigue** | 1.15 | 0.74 – 1.56 | <0.001 |
| **GAD-7** | 1.08 | 0.64 – 1.53 | <0.001 |
| **PHQ-9** | 1.01 | 0.54 – 1.47 | <0.001 |
| **PainDETECT** | 1.03 | 0.61 – 1.45 | <0.001 |
| **FM Severity** | 0.94 | 0.49 – 1.38 | <0.001 |
| **TJC–SJC diff** | 0.58 | 0.08 – 1.08 | 0.023 |
| rTPPT | –0.51 | –1.02 – 0.01 | 0.054 |
| CPM effect PDT | –0.49 | –0.97 – 0.00 | 0.051 |
| CPM effect PTT | –0.01 | –0.52 – 0.50 | 0.964 |
| TSP | 0.09 | –0.47 – 0.65 | 0.756 |

## Discussion

In this longitudinal study of patients with inflammatory arthritis followed from diagnosis, half of the patients continued to report clinically relevant pain at two years. Using a comprehensive patient assessment, including joint ultrasound we were able to obtain insights into both the inflammatory burden as well as sensory and psychological contributors, obtained by questionnaires and QST. These findings build on our observation that features of centrally mediated pain are present at diagnosis, and extend the, to date, very limited longitudinal data in early IA. We found that anti-inflammatory treatment reduced inflammation comparably across patients but that markers consistent with centrally mediated mechanisms and psychological distress, identifiable at diagnosis, were associated with pain persistence. In particular, 25% of patients had no improvement in their pain scores over the 24 month follow up.

By comprehensively examining inflammatory and non-inflammatory mechanisms from diagnosis, this study offers new insights into the relative contributions of these mechanisms to pain over time. We found that inflammatory markers (CRP, SJC, ultrasound scores, and PPT at inflamed joints) improve in almost all patients but that although patient reported outcome measures (PROMS) of centrally mediated pain (painDETECT, fibromyalgia criteria) and psychological distress improved on average alongside pain, some patients had no change. We used QST to expand on questionnaire data and potentially gain mechanistic insights into the causes of pain. The mean values of the QST markers of centrally mediated pain (trapezius (i.e. non-articular site) PPT, TSP, and CPM) remained largely unchanged. This may suggest that while DMARDs effectively reduce inflammation and may indirectly improve some centrally mediated symptoms (e.g., through mood, sleep, or enhanced perceived disease control), they do not directly and/or sufficiently alter central pain processing mechanisms as quantified by QST. In addition, this may reflect methodological limitations, as QST protocols, particularly the CPM paradigm, are known to have poor intersession reliability and lack definitions for clinically important changes[19–21].

Individuals with persistent pain had similar trajectories of improvement in swollen joint counts, ultrasound inflammation, and CRP following initiation of DMARD therapy as those without persistent pain. This pattern highlights a key message of our findings: pain persistence is not explained by inadequate control of inflammation. Instead, the separation between groups was defined by those with persistent pain having higher baseline and longitudinal scores for centrally mediated pain (fibromyalgia severity, painDETECT scores) and psychological distress. Our mixed-effects linear regression models, though requiring cautious interpretation due to sample size, also identified baseline PROMS of centrally mediated pain, but not inflammation, as predictors of subsequent persistent pain. Together, these findings suggest that while anti-inflammatory treatment reduces inflammation across patients, centrally mediated mechanisms primarily contribute to persistent pain and can be identified early in the disease course.

Comparison with existing literature is limited, as few longitudinal studies have followed patients from diagnosis or used the same patient-reported outcome measures, and most have focused on disease activity rather than pain specifically[4,22]. Using data from three large RA cohorts, including one in early IA (ERAN , BSRBR, and BSRBR-NB), McWilliams et al identified distinct pain trajectories (persistent pain, resolving pain ± low pain) based on SF-36 bodily pain scores. Across all cohorts, baseline predictors of persistent pain were higher health assessment questionnaire scores (HAQ) and a history of smoking. Another study used machine learning analysis of 789 patients followed for 5 years from diagnosis; it identified patient global assessment (PGA) and HAQ scores at three months as the strongest predictors of later persistent pain[22]. Other studies have longitudinally assessed patients with established IA. Those that use the same PROMS align with our findings in showing that higher painDETECT scores[23] and fibromyalgia symptom severity scores[24] as well as anxiety and depression are associated with worse disease activity and quality of life.

It is important to note that, although we used currently available tools to look for evidence of likely centrally mediated pain mechanisms in our cohort, some PROMS only reflect symptoms consistent with centrally mediated pain but do not provide any mechanistic insights. For example, painDETECT identifies neuropathic like pain and symptoms of pain sensitivity, which may be attributed to both centrally mediated pain and peripheral sensitisation. QST protocols aim to provide more detailed insights into underlying centrally mediated pain mechanisms but in our cohort baseline QST measures were not associated with persistent pain. The existing literature on QST as a predictor of long-term pain is limited, with only a small number of studies, generally inconclusive results, and a tendency to use treatment response as a proxy for pain outcomes. For example, low knee PPT in patients with low disease activity and abnormal CPM in those with high disease activity may predict poor treatment response[25], suggesting that QST markers may have limited clinical utility in predicting pain outcomes.

Nevertheless, overall, our findings suggest a potential role for central pain processing and possibly psychological distress from the outset, indicating either pre-existing centrally mediated pain or a vulnerability to long-term pain due to predisposing psychological and sensory factors. Given that we have shown that these markers can be plastic and modifiable, this highlights a potential window for early identification of patients at risk of persistent pain, and targeted intervention.

### Strengths and limitations

A strength of this study is the longitudinal design, with a comprehensive dataset collected from a real-world cohort of individuals with early inflammatory arthritis. In contrast to many registry-based studies which rely solely on routine clinical data, this study incorporated an in-depth assessment of pain drivers, including objective assessments of inflammation (MSKUS) and centrally mediated pain (static and dynamic QST) alongside clinical assessment and PROMs.

The cohort was representative of routine IA clinic populations, with 56% female participants and a diverse ethnic representation (only 55% identified as White British). Patients were recruited early in their disease process, with mean disease duration of 1.2 months. Follow-up rates were acceptable , with 92% of participants completing at least one follow up assessment. Participants underwent repeated assessments at multiple time points over a two-year period, allowing for detailed examination of pain and symptom trajectories. We used mixed-effects modelling to account for within-subject variability and incomplete data across timepoints and the factor-derived composite scores for inflammation and centrally mediated pain mechanisms avoided collinearity in subsequent modelling.

A limitation is that the sample size was 66. Although this exceeded the target sample size calculation for the original primary hypothesis, it limited statistical power for certain analyses, particularly multi-variable logistic regression analyses. To mitigate this, we performed linear regression with repeated measures to increase power and performed careful stepwise selection of variables. Whilst most participants completed follow up, some attrition did occur, leaving the possibility of responder or attrition bias. Finally, the inclusion criteria of a pain score ≥3 was selected based on prior literature and because some of our measures (e.g. painDETECT) required the presence of pain for assessment. To maintain consistency, and in line with previous studies that have used cut-offs between 2 and 4, we also applied this threshold when defining ‘clinically relevant persistent pain.’ However, this approach may limit the generalisability of our findings, as our sample may not fully represent the broader early inflammatory arthritis population, which includes patients with lower pain levels.

### Conclusion and Clinical implications

Half of participants continued to experience clinically meaningful pain two years after diagnosis, with 25% experiencing no pain improvement. This indicates a potential unmet need in pain management that may leave some patients feeling their symptoms are dismissed. Despite similar reductions in inflammation in all patients, centrally mediated mechanisms primarily contribute to pain persistence, reinforcing the need to move beyond an inflammation-only model of pain management, even in this early stage of IA. Encouragingly, features of likely centrally mediated pain such as fibromyalgia-like symptoms were not fixed and showed potential for improvement, suggesting they may be modifiable with targeted intervention. Baseline characteristics including higher painDETECT scores and psychological distress predicted persistent pain, offering an opportunity for early identification of patients at risk. These findings support the potential value of incorporating non-inflammatory pain assessment into early care. Future research should focus on validating stratification tools that include centrally mediated pain markers, identifying effective targeted interventions, and testing these in stratified trials. Longer-term follow-up will also be important to understand the trajectory of centrally mediated pain features over time.

## Data availability

The data underlying this article will be shared on reasonable request to the corresponding author.

## Patient and public involvement

Patients and the public were involved throughout this research, from shaping the research question at a workshop with expert patients, clinicians, and academics, where pain was identified as the top priority in inflammatory arthritis, to confirming its importance at a subsequent public engagement event. Patients contributed to study design, with National Rheumatoid Arthritis Society members reviewing pain questionnaires and two patient research partners (PRPs) refining the protocol, consent forms, and information materials. PRPs were also involved in interpreting findings, ensuring conclusions reflected patient perspectives, and providing feedback on manuscript wording; both (TE and RW) are included as PRP authors. Results have been shared with patients and the public through lay summaries and presentations at patient engagement events.

